# Epidemiologic, Clinical, and Biological Characteristics of Adult T-cell Leukemia / Lymphoma in Martinique (1983-2013)

**DOI:** 10.1101/2020.07.15.20126599

**Authors:** Plumelle Yves, Michel Stephane, Delaunay Christine, Kone Moumouni, Meniane Jean-Côme, Christian Derancourt, Merle Harold, Molinié Vincent, Panelatti Gérard

## Abstract

We report one of the largest non-Japanese cohorts of adult T-cell leukemia/lymphoma (ATL). A total of 175 cases were identified in Martinique between 1983 and 2013. The sex ratio was 1.01. The median age was 56 years. The overall incidence rate was 1.16/10^5^ inhabitants/year, with a risk 1.29 times higher for men. The distribution of clinical types (acute, lymphoma, and chronic) was 62.9%, 29.1%, and 8%, respectively. Median survival time was 3.06, 8.13, and 45.16 months, respectively, for the acute, lymphoma, and chronic types (p <0.001). Survival was significantly higher for lymphoma type with skin lesions (median 13.96 months versus 6.06, p <0.002) and for the acute type without hypercalcemia (5.1 versus 2.4 months, p <0.01). Symptoms associated with hypercalcemia present in 46.9% of patients and skin lesions in 43.4% were the best-performing clinical signs for the diagnosis of ATL. However, only 16.9% had these manifestations. *Strongyloïdes stercoralis* infection was documented in 42.5% of patients. Twenty-three patients had an atypical phenotype, including 14 cases with CD4-CD8- and 4 with CD4+CD8+. Twenty-four patients did not express CD25 with no significant impact on overall survival. The hyperploidy, trisomy of chromosome 3 and rearrangements of chromosome 14 were the most frequent karyotype abnormalities.

## Introduction

Adult T-cell leukemia/lymphoma (ATL) was first described in the Southwest islands of Japan in 1977 [1]. Then, the HTLV-1 (human T-lymphotropic virus-type 1) was isolated in the United States in 1981 in two patients, one with mycosis fungoïdes and the other with Sezary syndrome [2,3], which have been consequently renamed. Since then, this virus and ATL were identified in the Caribbean islands, Subtropical Africa, South America, and the Southeastern United States. HTLV-1 was also associated with various inflammatory diseases, including HAM/TSP (HTLV-1-associated myelopathy/tropical spastic paraparesis) [4-6] and infective dermatitis associated with HTLV-1 (IDH) [7,8]. The main clinical characteristics of ATL are lymphadenopathy, hepatosplenomegaly, skin lesions, and hypercalcemia. The criteria established by the Japanese Lymphoma Study Group/Shimoyama classification (LSG) recognizes 4 types of ATL: acute, lymphoma (aggressive forms), chronic,and smoldering (indolent forms), depending on the importance of lymphocytosis, tumoral syndrome, hypercalcemia, and level of lactate dehydrogenase (LDH) [9]. The neoplastic cells display a mature T-cell phenotype CD2+,CD3+,CD4+,CD7-. The ATL cell expresses constitutionally the IL2-receptor alpha-chain (CD25), regarded as a tumor marker of ATL. It has an intrinsic resistance to conventional chemotherapies. Chromosomal abnormalities are common and complex [10]. Infection with *Strongyloïdes stercoralis* (Ss) is common and defined as a risk factor [11-14]. All these factors contribute to a poor prognosis with a median survival time (MST) of 6.2, 10.2, and 24.3 months for acute, lymphoma, and chronic types, respectively [9].

Martinique (French West Indies) as an endemic area for HTLV-1 and ATL was recognized in 1983 [15-17]. Approximately 2% of the population tested positive for HTLV-1 [15,16]. The level of viral endemic was within the range of rates observed in the Caribbean [18] but lower than that observed in endemic areas of Japan [19]. However, the incidence rate of ATL among patients infected with HTLV-1 [20] was comparable to the one observed in Japan [21]. So far, epidemiologic and clinical data about ATL in Martinique were only partially reported [20,22,23]. In particular, partial data were published in a previous report [22] and reported in the present work. Here we proposed to study the epidemiologic, clinical, and biological characteristics of patients with ATL, identified over 30 years in Martinique, and to compare our experience with acquired data. In particular, we studied the immune-phenotype and karyotype of ATL cells.

## Patients and methods

### Patient population and diagnosis criteria

This retrospective analysis concerns ATL patients, extracted from a specialized registry of all hematologic malignancies of Martinique, identified between 1983 and 2013. Patients residing in Martinique for at least 6 months were retained. They were treated at the University Hospital Center of Martinique. All patients presented clinical features compatible with ATL disease and cytologic and / or histologic and / or immunologic confirmation and serologic confirmation of HTLV-1 infection. Clinical types were classified according to the strict criteria of LSG [9]. Thus, when rereading files, the lymphoma type was retained in the study only if the percentage of circulating atypical lymphocytes was strictly inferior or equal to 1% (% evaluated in relation to total lymphocytes). Consequently, patients with lymphocytosis less than 4.10^9^/L and atypical lymphocytes level more than 1% (oligo- leukemic pattern) were classified as leukemic type (and not smoldering) because they presented either lymph nodes, nodular skin lesions, or gastrointestinal impairment or pleural effusion or a rate of LDH greater than 1.5 times the normal upper limit. Patients presenting exclusively nodular skin lesions were classified as lymphoma or chronic type. Cytopenias were defined according to the WHO’s criteria: anemia if hemoglobin (Hb) <10g/dL and thrombocytopenia if platelets <100.10^9^/L. Because of the African origin of the population, neutropenia was noted if polynuclear neutrophils were <1.10^9^/L. Our retrospective observational study using existing data was exempt from requiring written informed consent in accordance with the declaration of Helsinki. So, approvalby the ethical committee was not necessary.

### ATL cell study methods

Immunophenotyping studies were performed on peripheral blood lymphocytes in 105 patients (94 acute, 11 chronic). Five cases were analyzed by fluorescence microscopy and the remainder by flow cytometry (FacsCalibur, Becton-Dickinson). The antibodies tested were CD2, CD3, CD4, CD5, CD8, TCR alpha-beta, CD7, CD25, HLA-DR, CD45RO, CD27, CD38, CD30. The positivity threshold was set at 30%. CD3 and CD7 were also analyzed in the cytoplasm in 10 and 15 patients, respectively. The immune phenotype was also performed in 18 skin biopsies, 26 lymph node biopsies, 24 bone marrow biopsies, and 13 broncho- alveolar lavages. The proliferation index was analyzed in cytology with Ki67 antibody and in histology with MIB1 antibody. The positivity threshold was set at 10%. In addition, monoclonal integration of HTLV-1 in tumor cells was investigated in 19 patients using inverse PCR or Southern blot and T-clonality in 5 patients. A karyotype analysis (performed by CERBA laboratory, 95310 Saint-Ouen-l’Aumône, France) was on circulating lymphocytes in 34 patients. Lymph nodes specimens (52 patients/pt), bone marrow cores biopsies (24 pt), and skin biopsies (19 pt) were submitted for routine histology.

### Statistical analysis

To estimate the incidence rates of ATL specific for gender and age, we use INSEE data (National Institute for Statistics and Economic Studies) for 1990 to 2013. For 1983 to 1989, extrapolations were made. Due to the rarity of cases aged under 20 years, the incidence rates were calculated for a population older than 20 years old. Overall survival was calculated from the date of diagnosis to the date of death or of last news. Patients lost to follow-up were considered at risk until the date of last contact, at which point they were censored. Survival curves were established using the Kaplan-Meyer method based on the following parameters: type of ATL, skin lesions, hypercalcemia, typical/atypical phenotype, and intergroup survival was compared using the log-rank test. The MST was defined as 50% on the survival curves. The Cox regression model was used to evaluate the prognostic impact of the following factors: age (<or> 40 years), sex, type of ATL, tumoral syndrome, calcemia, LDH level (<or> 2N), eosinophilia (> 1.10^9^/L), and Ss. The chi-2 test was used for analysis of distribution of patients among the various types of ATL according to the same variables. The Fisher exact test was used when the distribution analysis concerned only the acute and lymphoma types or aggressive and indolent forms.The Student test was used to analyze changes in the age at diagnosis. The chi-2 test was used to test the independence of hypercalcemia and skin lesions. Significance was achieved at p <0.05. Statistical analysis was performed using SAS.9.2 software (SAS Institute, Inc., Cary, NC, USA).

## Results

### Epidemiologic data

During the study period, Martinique’s population increased from 364,000 to 381,000 inhabitants, consisting of 96% of people of mixed African-Caucasian descent. The main problem concerning population-based registries is the lack of completeness. In Martinique, the risk of incompleteness is limited because there is only one laboratory that specializes in cytology and one clinical hematology unit on the island. Therefore, the registry included all patients referred to the hospital. A total of 175 new cases of ATL were identified. All patients were of mixed African Caucasian descent. Seven patients were lost to follow-up. There were 88 men and 87 women. Thirty-seven patients (21%) from 16 families were affected in the context of familial ATL, affecting siblings or mother and child. The median age was 56 years for each sex (range 16-95). Twenty-nine patients (16.6%) were under 40 years old, and 18 (10.3%) were over 80. Standardized incidence rates per 100000 inhabitants/year are shown in figure 1. In particular, the overall standardized incidence was 1.16/10^5^ inhabitants/year with an ATL risk 1.29 times higher for men than for women. The specific incidence rate of age increased up to the 40-49 years age group in both sexes.

**Figure 1:**
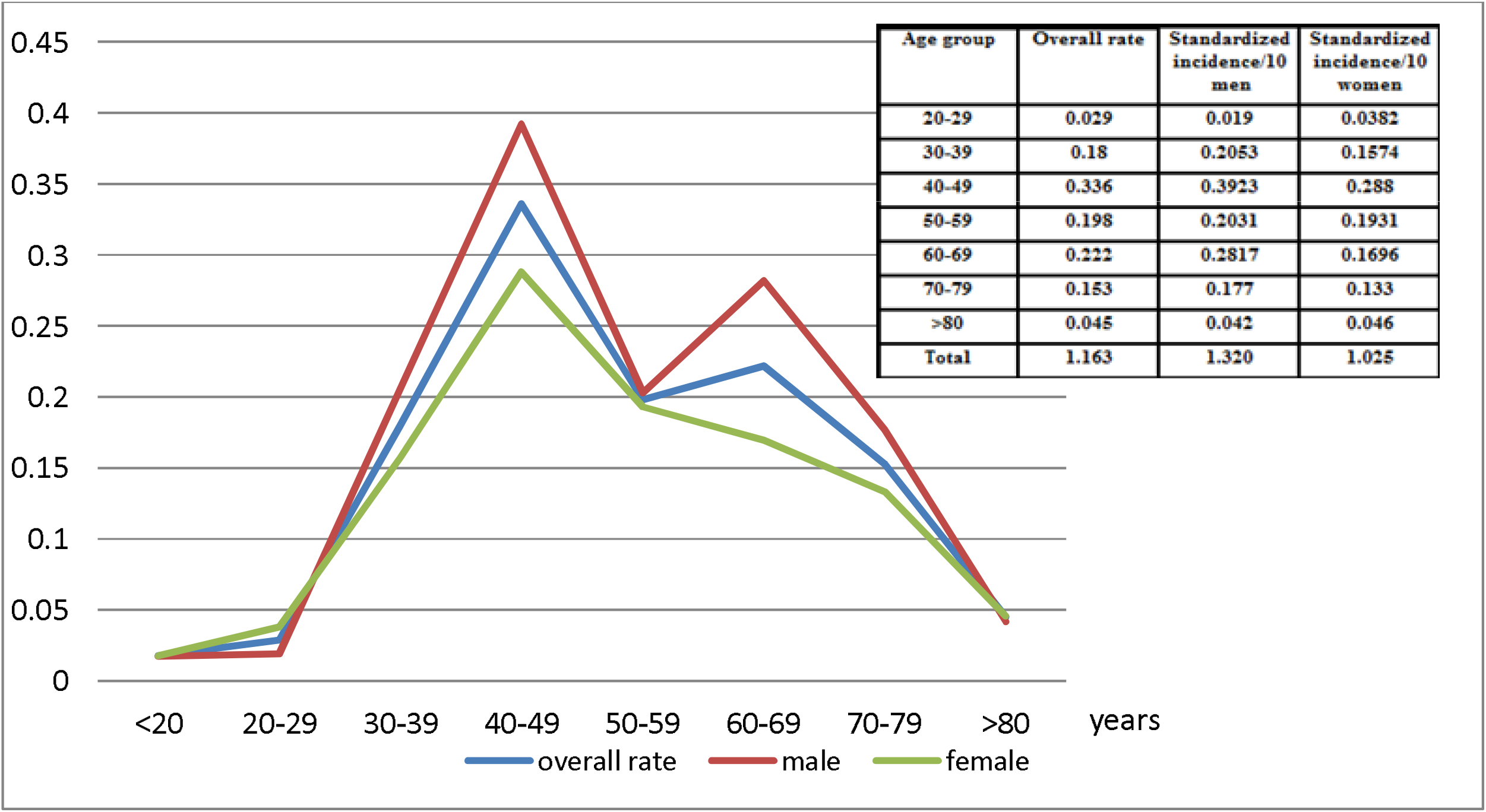
Age standardized incidence rates of ATL in Martinique from 1983 to 2013 among the adult population per 100000 inhabitants per year.

The distribution of clinic types and MST are shown in figure 2. The MST of the 3 patients with exclusive nodular skin lesions was 23, 52, and > 150 months, respectively. Thirty patients (17.1%) had a survival of less than 1 month. Ten patients (5.7%) (median age: 45 years old, 5 lymphoma type and 5 chronic type, 8 with skin lesions) that we reported in an earlier work [22] showed a survival greater than 10 years. Survival was significantly longer for the acute type without hypercalcemia (5.1 versus 2.4 months, p <0.01) (Figure 3) and the lymphoma type with cutaneous lesions (median 13.96 months versus 6.06, p <0.002) (Figure 4). The Cox regression model showed, in general, a significant negative impact of hypercalcemia on survival at 3, 6, and 12 months (p <0.001) and a decreased survival for the acute type compared to lymphoma type at the same times (p <0.001) (Table I).

**Table I.** Survival analysis of ATL patients depending on the ATL type, age and biological and clinical variables (Survival factors, Cox univariate model).

|  |  | Survival to 3 months | Survival to 6 months | Survival to 12 months |
| --- | --- | --- | --- | --- |
| Variables |  | RR [95%CI] | RR [95%CI] | RR [95%CI] |
| Type | Lymphoma | 1 (ref.) | 1 (ref.) | 1 (ref.) |
|  | Acute ATL | 0,36** [0,19 - 0,68] | 0,39** [0,23 - 0,65] | 0,51** [0,34 - 0,76] |
| Age | =<40 ans | 1 (ref.) | 1 (ref.) | 1 (ref.) |
|  | > 40 ans | 0,96 [0,87 - 1,06] | 0,94 [0,86 - 1,03] | 0,92* [0,86 - 0,99] |
| ADP | negative | 1 (ref.) | 1 (ref.) | 1 (ref.) |
|  | positive | 0,96 [0,88 - 1,04] | 0,93 [0,86 - 1,00] | 0,94 [0,88 - 1,00] |
| Ca | negative | 1 (ref.) | 1 (ref.) | 1 (ref.) |
|  | positive | 0,87** [0,82 - 0,93] | 0,91** [0,87 - 0,96] | 0,93* [0,89 - 0,97] |
| HM | negative | 1 (ref.) | 1 (ref.) | 1 (ref.) |
|  | positive | 0,99 [0,92 - 1,06] | 0,94* [0,89 - 0,99] | 0,96 [0,92 - 1,02] |
| CL | negative | 1 (ref.) | 1 (ref.) | 1 (ref.) |
|  | positive | 1,06 [0,99 - 1,13] | 1,05 [0,99 - 1,11] | 1,06* [1,01 - 1,11] |
| LDH | negative | 1 (ref.) | 1 (ref.) | 1 (ref.) |
|  | positive | 0,95 [0,89 - 1,01] | 0,96 [0,91 - 1,01] | 0,96 [0,91 - 1,00] |
| SM | negative | 1 (ref.) | 1 (ref.) | 1 (ref.) |
|  | positive | 0,97 [0,90 - 1,04] | 0,95 [0,90 - 1,01] | 0,97 [0,91 - 1,02] |
| Ss | negative | 1 (ref.) | 1 (ref.) | 1 (ref.) |
|  | positive | 1,02 [0,95 - 1,10] | 1,02 [0,95 - 1,08] | 1,02 [0,96 - 1,08] |
| HE | negative | 1 (ref.) | 1 (ref.) | 1 (ref.) |
|  | positif | 1,00 [0,93 - 1,09] | 1,02 [0,95 - 1,09] | 1,01 [0,95 - 1,07] |

RR, relative risk, \*significant p value<0.05, \*\*significant p value <0.001, CI: confidence interval, ADP: adenopathy, HM: hepatomegaly, SM: splenomegaly, CL: cutaneous lesions, Ss: *Strongyloides stercoralis*, HE: eosinophilia > 1.10<sup>9</sup>/L, Ca: hypercalcemia, LDH: lactate deshydrogenase greater than two times the normal upper limit.

**Figure 2:**
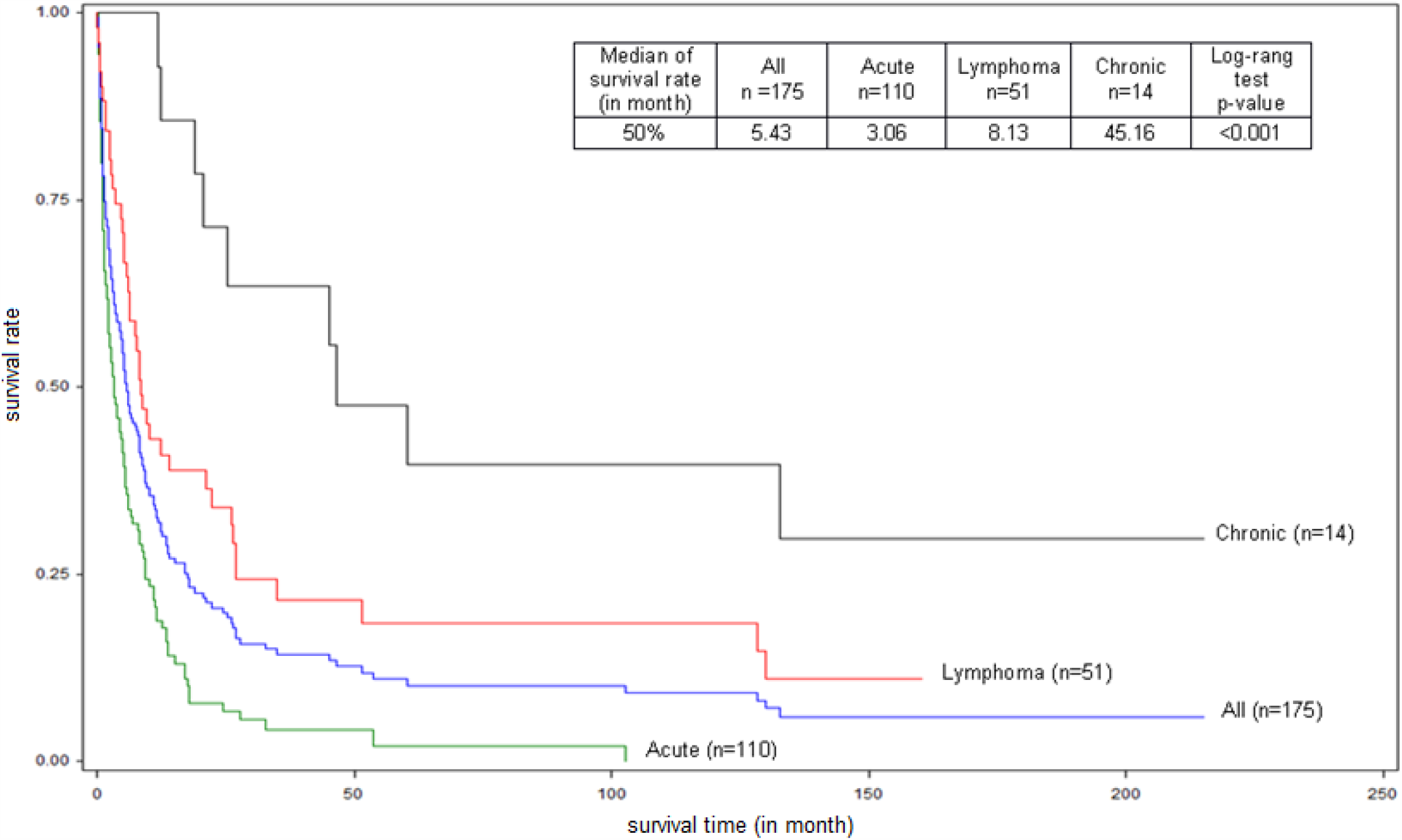
Global and specific survival, by clinical subtypes (acute, chronic, lymphoma), of Adult T-cell Leukemia/Lymphoma (ATL) patients diagnosed between 1st January 1983 and 31st March 2013 in Martinique. (N=175)

**Figure 3:**
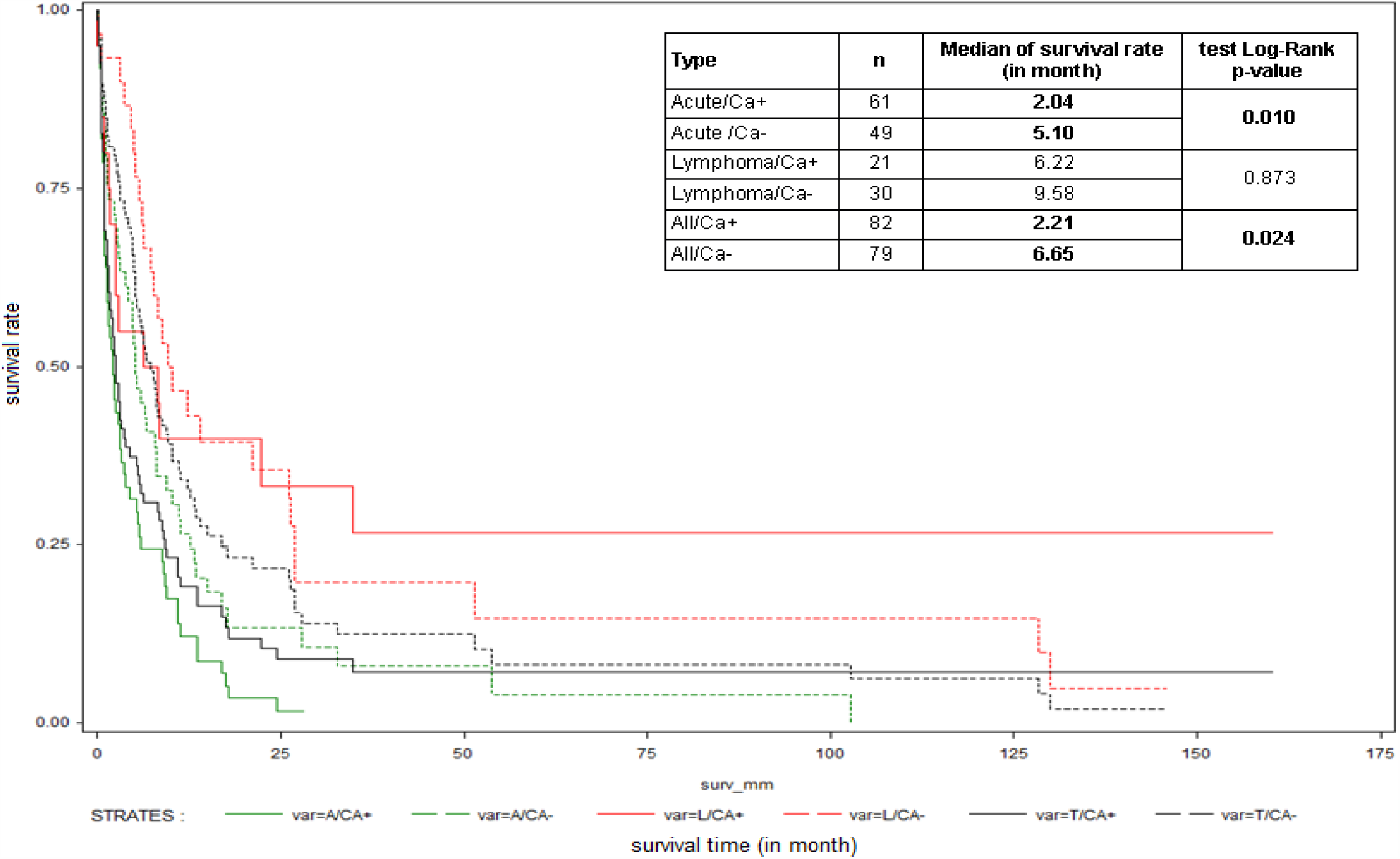
Global and subtype-specific survival (acute, lymphoma), according to presence of calcemia (Ca+) or absence of calcemia (Ca-), of Adult T-cell Leukemia/Lymphoma (ATL) patients diagnosed between 1st January 1983 and 31st March 2013 in Martinique.

**Figure 4:**
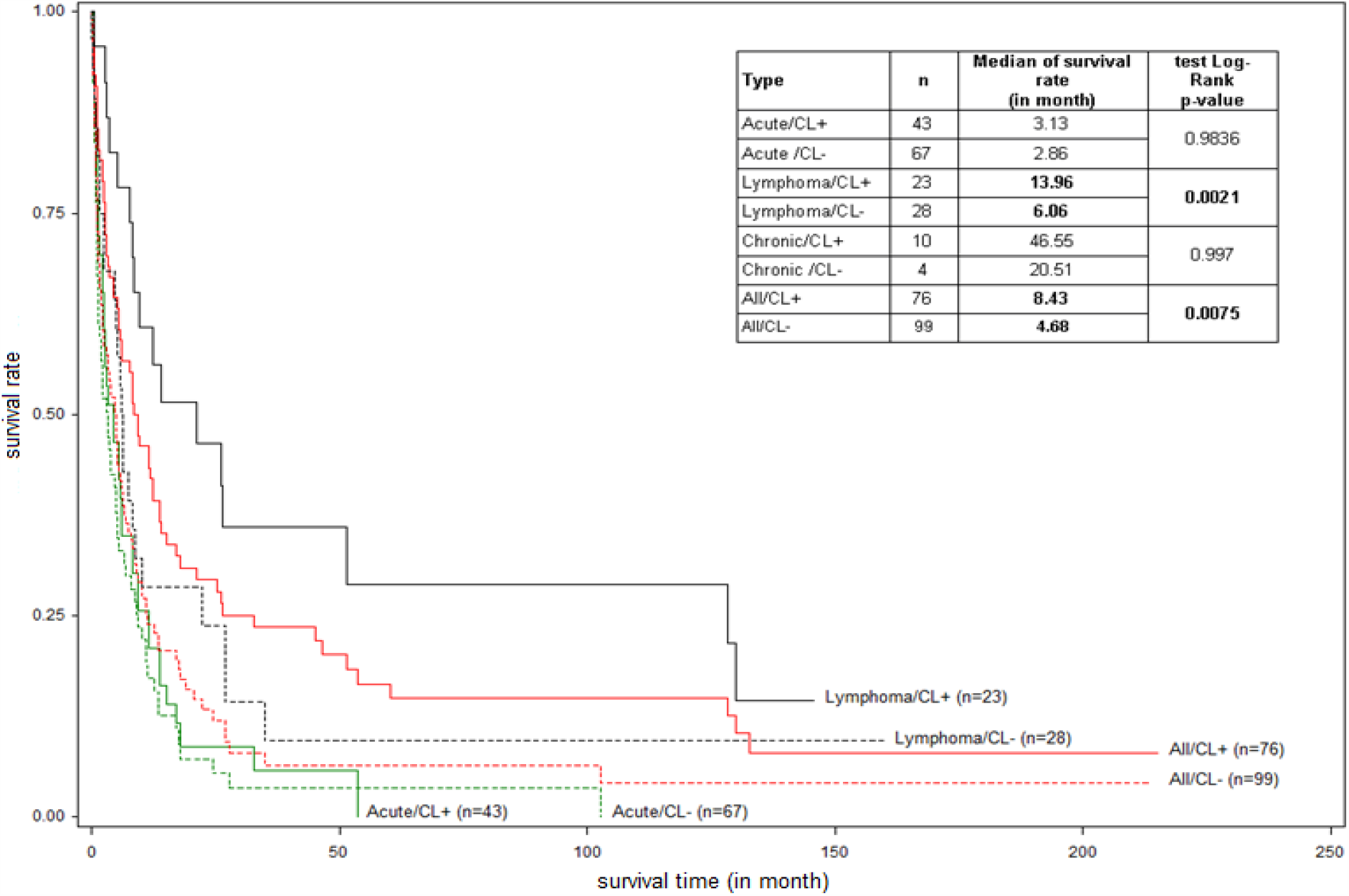
Global and subtype-specific survival (acute, lymphoma, chronic), according to presence of cutaneous lesions (CL+) or absence of cutaneous lesion (CL-), of Adult T-cell Leukemia/Lymphoma (ATL) patients diagnosed between 1st January 1983 and 31st March 2013 in Martinique. Because of the small number of patients, the survival curves of chronic subtype are not drawn.

### Clinical and biological features

Concerning the treatment, in brief, lymphoma type received an initial induction of chemotherapy, and leukemic types (acute and chronic) received the combination of interferon-alpha and antinucleoside (zidovudine or zalcitabine). Treatment for strongyloidiasis was performed in all patients.

Two patients had eczema during childhood, which disappeared at puberty. Four patients had HAM/TSP before ATL. Eleven patients had autoimmune diseases: systemic lupus erythematosus (3 patients), antiphospholipid syndrome (1 patient, 13 years before ATL), Sjögren syndrome (5 patients), scleroderma (1 patient), and sarcoïdosis (1 patient, 3 years before ATL) [24]. Moreover, 3 cases of *a frigore facial palsy* were observed.

Clinical and laboratory data are presented in table II. In particular, the symptoms associated with hypercalcemia and skin lesions were the most reliable clinical signs for the diagnosis of ATL. Patients with only hypercalcemia, those with only skin lesions, and those with both represented 30.8 %, 27.3 %, and 16.9 %, respectively. However, the two variables were not significantly independent (p >0.05). Hypercalcemia was significantly more frequent in the acute type than in the lymphoma type (p = 0.001). Erythema, papules, nodules, and ichthyosis-keratosis were observed in 41%, 30%, 26%, and 19% of patients, respectively. The frequency of skin lesions was significantly lower in aggressive forms than in indolent forms (66/161 versus 10/14, p = 0.027). In 4 patients, bone marrow was infiltrated by ATL cells.

Two patients had subretinal tumor infiltration; one progressed to blindness [25]. Three patients had acute pancreatitis: two had acute ATL, and one had chronic ATL. One patient with acute ATL showed an enlarged thymus, and another developed an acute ATL during pregnancy.

**Table II.** Distribution of age, gender and clinical and biological signs - expressed as %(number of patients) - according to patients groups (all, acute, lymphoma and chronic type) (Fisher exact test or Chi-2 depending on variables, Student test for age, significant p< 0.05)

| Variables | All (n=175)<br>% (N) | Acute(n=110)<br>% (N) | Lymphoma(n=51)<br>% (N) | Chronic(n=14)<br>% (N) | P value |
| --- | --- | --- | --- | --- | --- |
| Age± SD (years) | 56.92 ± 16.96 | 57.82±17.24 | 56.34 ± 16.13 | 51.9 ± 17.89 | 0.640 |
| SEX | 88M/87F | 56M/54F | 25M/26F | 7M/7F | 0.467 |
| ADP | 80.0 (140) | 83.8 (92) | 78 (40) | 57.1(8) | **0.004** |
| HM | 21.7 (38) | 25.2 (28) | 18 (9) | 7.1 (1) | 0.160 |
| SM | 18.3 (32) | 22.5 (25) | 12.0 (6) | 7.1 (1) | 0.108 |
| CL | 43.4 (76) | 39.1 (43) | 45.0 (23) | 71.4 (10) | **0.017** |
| APS | 42.3 (74) | 47.3 (52) | 35.3 (18) | 28.6 (4) | 0.609 |
| WL | 24.6 (43) | 34.5 (38) | 9.9 (5) | 0 | **0.003\*** |
| OL** | 15.4 (27) | 14.5 (17) | 19.6 (10) | 0 | 0.406* |
| CNS/CF+*** | 8.0 (14) | 11.8 (13) | 2 (1) | 0 | 0.054* |
| PE | 12.6 (22) | 10.9 (12) | 19.6 (10) | 0 | 0.253* |
| Ascitis | 4.6 (8) | 5.5 (6) | 3.9 (2) | 0 | 0.112* |
| Ss**** | 42.5 (51) | 39.0 (30) | 45.5 (15) | 60.0 (6) | 0.690 |
| Ca | 46.9 (82) | 55.5 (61) | 41.2 (21) | 0 | 0.062* |
| HE | 16.0 (28) | 18.9 (21) | 10.0 (5) | 14.3(2) | 0.395 |
| LDH | 55.4 (97) | 70.0 (77) | 40.0 (20) | 0 | **0.001\*** |
| Hb<10g/dL | 10.5 (18) | 6.4 (7) | 21 (10) | 7.1(1) | <0.01 |
| Platelets<100.10<sup>9</sup>/L | 10 (17) | 12.8 (14) | 4.2 (2) | 8.3 (1) | NS |
| PNN<1.10<sup>9</sup>/L | 3.6 (6) | 1.2 (2) | 4.3 (2) | 14.3 (2) | NS |
| PNN>10.10<sup>9</sup>/L | 20 (34) | 25.9 (28) | 8.5 (4) | 14.3 (2) | <0.02 |

ADP: adenopathy, HM: hepatomegaly, SM: splenomegaly, CL: cutaneous lesions, APS: abdominal pain syndrome, WL: weight loss, OL: osteolytic lesions, CNS/CSF+: central nervous system/cerebrospinal fluid + (ATL cell presence in 11 patients), PE: pleural effusion, Ss: *Strongyloides stercoralis*, Ca: hypercalcemia, HE: eosinophilia $> 1.10^9/L$ , LDH: lactate deshydrogenase greater than two times the normal upper limit, SD: standart deviation.
\* p is calculated by Fisher exact test, \*\* skull osseous lacuna in 11 patients, \*\*\* presence of atypical cells in 5 patients, \*\*\*\* 51/120 tested (acute: 77, lymphoma: 33, chronic: 10)

Infectious complications with gram-negative bacilli or as gastrointestinal candidiasis were common in medical history, diagnostic period, and terminal phase. In particular, Ss infection was documented in 42.5% (51/120) of the tested patients and preceded the diagnosis of ATL by 1.5 to 27 years. Ten cases of disseminated strongyloidiasis were accompanied by septicemia, meningitis, or pneumonia with gram-negative bacilli. Four cases of *Pneumocystis jirovecii* infection were documented. Two patients were co-infected with HIV.

During follow up, 154 patients (104 acute type, 41 lymphoma, chronic 9) died, with an overall MST of 4.68 months, 2.86 for acute type, 6.22 for lymphoma type, and 48 for chronic type. Hypercalcemia was the most constant biological (33%) sign and the leading cause of death. Absent at diagnostic period, it arose during the evolution in 25 patients. Thus, 61% (107/175) of patients experienced hypercalcemia during ATL. Two patients died of acute pancreatitis, and 2 died of macrophagic activation syndrome. Seventeen patients died due to infection. The secondary appearance or spread of skin lesions was observed in 4 and 2 patients, respectively. No strict lymphoma progressed to a leukemic form. There were eight oligo-leukemic cases: 4 were evolving towards a clear leukemic state; the other four retained the oligo-leukemic profile until death. During evolution, lymphopenia of less than 0.5.10^9^/L or 0.2.10^9^/L was noted in 34 (22%) and 17 patients, respectively.

### Cytology-histology, phenotype, and karyotype

In leukemic forms (70.9%, 124/175), atypical cells showed a net cellular pleomorphism, nuclear irregularities, and dense and blistered chromatin, conferring the status of “flower cell,” sometimes Sezary-like cell. The degree of nuclear irregularities was not related to the clinical type (unshown). In 79 cases (36 lymphomas and 43 leukemia forms), the pleomorphic character was observed mainly on skin or lymph node histology. Tumor infiltration was always diffuse. In 6 cases, Reed-Sternberg-like cells were described.

In most cases, the ATL cell showed the post-thymic activated T-phenotype, CD2, CD3, CD4, and CD25. The low expression of CD3 and alpha-beta TCR was observed in all cases analyzed. The absence of CD7 membrane expression was observed in 75/96 cases tested as well as in the cytoplasm in 15 cases tested. The activation markers CD45RO and CD27 were always expressed; CD38 in 51.2% (22/42 tested) and HLA-DR in 27.6% (16/58 tested). This typical phenotype concerned 81% of leukemia patients in our series. However, 23 patients (20 with leukemia/19% and 3 with lymphoma) showed a different phenotype (Table III). The survival of patients with an atypical phenotype significantly differed from patients with a typical phenotype (median: 2 months versus 5.76, p = 0.013) (Figure 5). In addition, 24/107 tested (22.4%) (17 acute, 4 lymphoma, chronic 3) did not express CD25, with no significant impact on overall survival (4.97 versus 5.1 months, p = 0.823) (Figure 6). The expression of CD7 had no significant impact on overall survival (2.25 versus 3.75 months, p>0.05). The measured proliferation index was moderate to strong in 17/19 patients tested.

**Table III.** Patients with atypical phenotype.

| Atypical phenotypes | Patients (N=23) |
| --- | --- |
| **CD2-CD4+** | **1** |
| **CD4-CD8-** | **14** |
| CD3+CD4-CD8- | 7 |
| CD2-CD4-CD8- | 2 |
| CD7+CD4-CD8- | 5 |
| **CD4+CD8+** | **4** |
| CD3+CD4+CD8+ | 2 |
| CD2-CD4+CD8+ | 1 |
| CD3-CD4+CD8+ | 1 |
| **CD3-** | **4** |
| CD3-CD4+ | 2 |
| CD3-CD3cyt+ | 1 |
| CD3-CD8+ | 1 |
| **CD25-** | 24/107 (22.4%) |
| **CD7+** | 21/96 (21.9%) |

**Figure 5:**
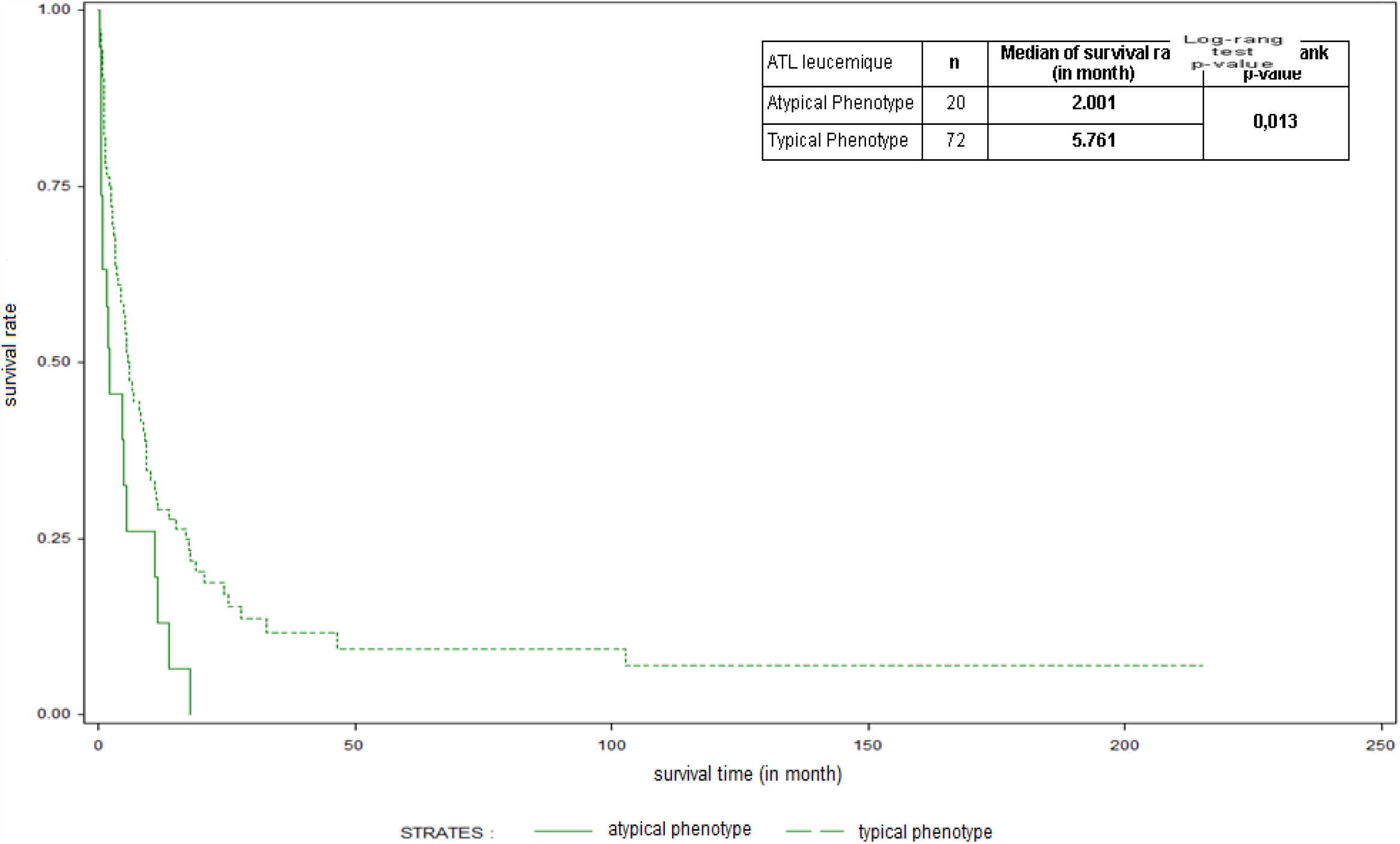
Survival curves of patients with ATL according to typical and atypical phenotypes.

**Figure 6:**
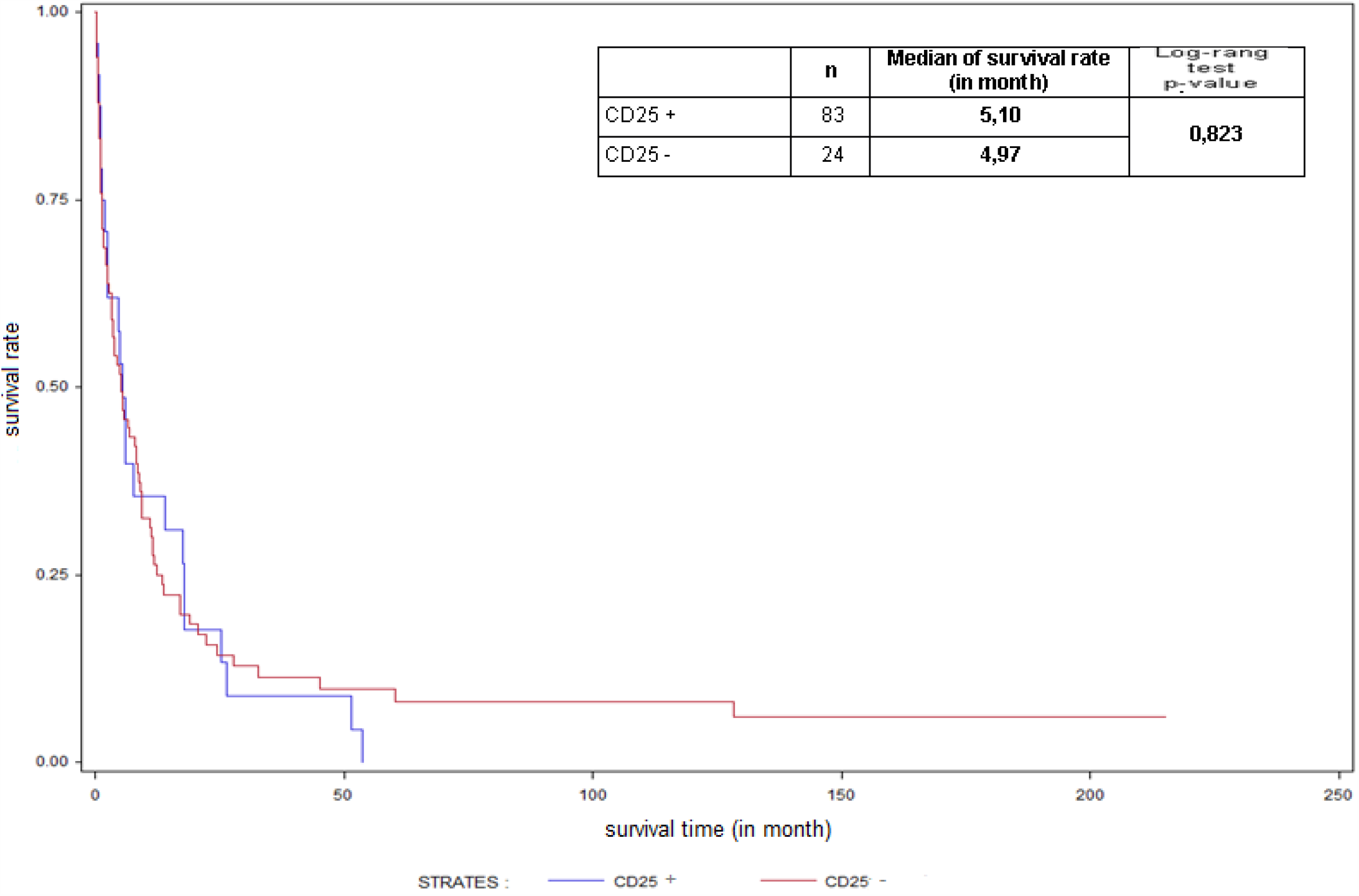
Survival curves of patients with ATL according to the expression of CD25.

At the karyotype level, the most common abnormalities were hyperploidy (chromosomes > 50) (26.5%, 9/34, 2 with long survival), trisomy 3 (35.3%, 12/34) isolated or associated with rearrangements including translocations, and trisomy 7 and 18 (20.6%, 7 patients each). Chromosome 14 was involved in rearrangements (14.7%, 5 patients) involving q32 or q31 loci, as well as a deletion of the 14q11 locus, one monosomy, and two trisomy. The most common monosomies involved chromosomes 10, 18, and 20 (4 times each) and chromosomes 2, 3, 11, 13, 15, and X (3 times each). Supernumerary marker chromosomes were observed in 30% (10/34) of patients. The presence of 2 or more clones and clonal evolution was documented in 12 (35.3%) and 6 patients, respectively. Abnormalities were not related to the clinical aspects (unshown).

## Discussion

### Epidemiologic data

Martinique is a high endemic area for HTLV-1 and ATL [15,16,20,22,23]. The higher risk among men of our cohort (figure 1) confirmed data from Japan [26]. Because of the severity of disease (many survivals of less than one month), the incidence rate should be interpreted as minimum. Moreover, the improvement of ATL incidence data requires analyzing proviral and TCR-rearrangement clonality for all cases of T-lymphoma to rule out fortuitous HTLV-1+ cases with no causal relationship to the ATL. In our series, the frequency of familial cases was higher than that reported by Japanese teams (21% versus 2.1%) [26]. When a patient develops ATL, the risk for one of his siblings also developing ATL increases, suggesting a common ground more favorable to ATL tumorigenesis than in HTLV-1 families without ATL cases. However, age of onset, initial clinical presentation, and evolution (overall survival) differed from one member of the same family to another, as evidenced, in particular, by the two families in which one in two patients had a long survival [22]. This suggested that changes in the environment, endogenous or exogenous, specific to each individual would promote the tumoral future of HTLV-1 cell clones: deleterious geographical environment (pesticides), infectious context of perinatal period or after the HTLV-1 infection such as Ss infection, or thymic different status. Our data on age, sex ratio, proportion of patients with risk factors such as age above 40 years, hypercalcemia, high LDH rate, tumoral syndrome, and the MST of different types (table II, figure 2) were comparable to those of Japan and Jamaica [9,27]. Consistently with the literature, we could not find a correlation between the type of ATL and age of onset (table II) [9]. Regarding the median age, unlike a first report [20], we could not find any difference with Japan. However, the number of patients under 40 years old was significantly higher than in the Japanese series (16.6% versus 7.1%, p = 0.004) [9]. Our series confirmed the rarity of the ATL in adolescents [28].

### General biological and clinical data

In our series, hypercalcemia was a discriminating factor of severity consistent with the literature [9], especially in the acute type (figure 3). Its permanence or its appearance signified the therapeutic inefficiency. Like others, we observed pancreatitis: the association of hypercalcemia, or chronic alcoholism, and / or tumor infiltration could be determining factors [29]. The high frequency of patients with skin lesions confirmed the literature [9,30-32]. Clinical research of skin lesions is an important step in the diagnosis of ATL. The presence of skin lesions contributes to the classification of ATL, since the smoldering type is defined by this single clinical sign [9], and a strict cutaneous form is recognized [33,34]. However, the exclusive skin involvement and the emergence or extension of skin lesions during the evolution were rare. Their presence and clinical characteristics could have a prognostic impact [30-32]. So, they were less frequent in aggressive forms than in indolent forms. Regardless of the type of ATL, the presence of skin lesions was associated with better survival, especially for the lymphoma type (figure 4). In particular, their high frequency associated with a younger age was noted in patients with a long survival [22]. However, the comparison of survivals with other series was difficult because of their small size, different types of ATL recruitment, including high frequency of smoldering type; diversity of histologic characteristics of skin lesions, and their primary or secondary nature [30-32]. Moreover, although the skin lesions or hypercalcemia were very common, simultaneous frequency of both was low in our series and observed by another [26], suggesting the existence of different ATL clones. All these observations suggested that: (i) the skin tissue was a primary, early, and privileged location of the ATL cell and that the infiltration of other tissues was secondary and metastatic in nature, and (ii) early treatment of skin lesions could slow the evolution toward a more aggressive form. This hypothesis is consistent with clonal succession [35,36] underpinned by the observation, in our series and others [10], of the simultaneous or successive presence of several clones and of the clonal evolution often mentioned in the karyotypes reports.

In our study, we have not identified IDH during the ATL patients’ childhood [7,8]. Thus, our series could not confirm IDH as an ATL risk factor in adults, as observed in Jamaica and Brazil [37,38]. The ocular tumor involvement, mainly subretinal, observed in aggressive ATL forms, is rare [24,39-41]. Its severity implies its systematic search, not only in all ATL patients, but also in HTLV-1 patients at risk of developing ATL—that is, ATL patients’relatives and HTLV-1 healthy carriers.

No involvement of the mediastinum or thymoma was observed, in accordance with the original description of the ATL in 1977 [1]. However, one patient presented a thymic hypertrophy. This observation may perhaps be linked to benign epithelial thymoma developed in a rat model carrying the HTLV-1 pX region [42,43] suggesting, in this patient, an unusual intrathymic overexpression of HTLV-1/TAX protein, which induces benign hypertrophy of the thymus.

Infection with Ss is common and defined as a risk factor for leukemogenesis in ATL [11-14]. This infection, diagnosed many years before ATL, was frequent in our series. But the number of overwhelming strongyloidiasis was relatively low, despite the persistent nature of the infection and the immunosuppression induced by HTLV-1. Furthermore, *Pneumocystis jirovecii* infection, reported in the literature [44], was rare in our series.

The blood count’s parameters and bone marrow were minimally affected in our series (table II), suggesting the weak involvement of the bone marrow in ATL. Several studies explained the good conservation of hematopoiesis: (i) the demonstration of an orientation of hematopoiesis to the T-cell lines, induced by HTLV-1 infection by altering expression of the microRNA associated with hematopoiesis [45], (ii) the absence of infection by HTLV-1 of hematopoietic progenitors from ATL patients [46], (iii) the transcriptional activity of TAX, observed *in vitro* on different genes stimulating hematopoiesis [47,48]. In practice, the infiltration of the bone marrow by tumor cells, sometimes observed, was to be interpreted according to the importance of circulating atypical lymphocytosis (peripheral blood contamination). Neutrophilia was common and often linked to an infectious or inflammatory disease. However, HTLV-1 (via the transcriptional activity of TAX *in vitro* observed on the *GMCSF* gene) could induce *de novo* or amplify the expression of this gene [47]. In addition, high levels of certain microRNAs, such as miR-223, observed in ATL cells, *ex vivo*, would encourage granulopoïesis [49]. These activities could explain the rarity of neutropenia throughout evolution. In critical phase (after chemotherapy), bicytopenia (anemia and thrombocytopenia) was frequent but moderate. In case of deep cytopenia, it is important to look for other causes, such as macrophage activation syndrome.

The ATL cell presents such characteristics that it is sufficient, in most cases of leukemia, to make the diagnosis. However, like others, we described Reed-Sternberg-like cells [50] to confront the context of hypercalcemia. Furthermore, due to atypical cells that can be few, researchers should be careful, especially in cases in which the lymphocytosis is less than 4.10^9^/L, and confront the analysis by flow cytometry [51]. This analysis is necessary for proper classification and fundamental for inclusion in clinical trials.

### LSG classification

Ranking the different types of ATL is an important step toward diagnosis due to the consequent therapeutic decision and the significance of its prognosis [52,53].The distribution of the types—acute, chronic, and lymphoma—was comparable with the distribution of the Japanese series [9]. However, in Martinique, unlike in Japan and Jamaica, we have not clearly identified any smoldering type: inefficient screening, rapid transformation to other more aggressive forms as suggested by a recent study [54]. Furthermore, the observation of a small percentage of circulating atypical cells associated with the detection of proviral and TCR rearrangement monoclonalities demonstrates the arbitrariness of the 1% threshold to distinguish the leukemic type from the lymphoma type [55]. The detection of many clones in the peripheral blood of patients classified as having lymphoma, without atypical cells circulating or monoclonality by southern blotting, emphasized that leukemia assessment was technology dependent [55].

In our series, some patients showed a long survival, including lymphoma type, which suggested that this aggressive clinical form did not prevent an indolent evolution [22]. Conversely, several reports showed in indolent forms a faster progression than expected, suggesting the need for careful clinical monitoring of these forms and the development of appropriate treatments [30,54]. Furthermore, a 20-year follow-up of 50 HTLV-1 carriers, called “pre-ATL” (defined as leukemic ATL without other organ involvement, hypercalcemia, and high LDH value that is almost identical to chronic/smoldering ATL) due to a monoclonal proliferation of T-cells, revealed a 42% cumulative probability of developing ATL [56], highlighting the difficult distinction between “pre-ATL” state and the smoldering and chronic types. Another study showed, for aggressive ATL, heterogeneous remission characteristics induced by chemotherapy [57]. These observations and the absence of clearly identified smoldering form in our series suggest that smoldering form is probably rarer than originally described. These facts demonstrated that the useful classification of LSG [9] during the diagnostic period was not satisfactory in terms of prognosis and insufficient for the choice of treatment, requiring the identification of new criteria. So, recently, the chronic form began to be classified into favorable and unfavorable subtypes based in the levels of LDH, urea nitrogen, and albumin [58].

The LSG classification raises the possibility of an evolution from one type to another [9]. However, in our series, no lymphoma type evolved into a leukemic form. This suggested that the natural history of tumor cells of each ATL type was different. This hypothesis is confirmed by the identification of specific genomic alterations of acute or lymphoma type that would develop through distinct genetic pathways or from different phenotype cells [59].

### Karyotype abnormalities

Because of the few patients analyzed, we were not able to demonstrate a link between survival or clinical aspects and an abnormal karyotype. However, the presence of hyperploidy in two patients with long survival suggested its non involvement in the prognosis [22]. *A*.*contrario*, our observation and others’, of a high frequency of abnormalities affecting chromosome 3 in the form of trisomy and translocations, and chromosome 14, in the form of translocations or deletions involving 14q11, 14q32, and 14q31 loci, suggesting a role for these abnormalities in the natural history of the ATL cell [10,60-63]. So, the analysis of Japanese Karyotype Review Committee’s report on aggressive ATL forms revealed that in 77% (62/81) of patients, chromosomes 3 and / or 14 were involved in abnormalities. Among them were 2 cases of aggressive ATL, but one indolent had a translocation involving both chromosome 3 and 14 [10]. However, 23% of patients in this report showed no abnormalities in chromosomes 3 and 14. Supernumerary marker chromosomes often observed in this report (52%, 42/91), as in our series, would host rearrangements involving these same loci. Other cutaneous T cell lymphomas, such as Sezary syndrome and mycosis fungoïdes, have karyotypes whose complexity is comparable to that of ATL [64-66]. However, the near absence of recurrent implications of these chromosomes reinforces the hypothesis of a specific role for abnormalities of 3 and 14 in ATL. Consistently with this view, one work demonstrated a correlation between the proliferation of ATL cells, the formation of “flower cell,” and overexpression of PI3K-AKT (phosphatidylinositol 3-kinase), induced by the alteration of the gene PIK3CB2 located at the 3q22 locus level [67]. Another recent work revealed that the constitutional activation of the PI3K-AKT signaling pathway in ATL cells resulted from the loss of expression of the tumor suppressor gene, *NDRG2* (N-myc downstream-regulated gene 2), located in the 14q11 region [68].

So, the great diversity of karyotypes observed in our study and others [10,60-63], often complex but sometimes almost normal, suggested that the majority of abnormalities are acquired secondarily to the pre-ATL cell and necessary for the survival of the completed ATL cell but without impact on survival.

### Phenotype and origin of the ATL cell

ATL cells in the majority of our patients presented a post-thymic mature T-phenotype expressing CD2, CD3, CD4 [69] (peripheral CD4+ T-cells as normal counterpart according to WHO classification 2008). The expression of CD25, a biomarker of the prototype ATL cell, whose constitutionality is admitted, was absent in 24 patients. Its expression was correlated neither to clinical type nor to survival (figure 6), suggesting a modest or passenger role in ATL leukemogenesis. Consistent with this view, in a recent trial of treatment with an antibody anti-CD25 (daclizumab), no responses were observed in 18 patients with ATL aggressive forms and only 6/16 partial responses in ATL indolent forms [70]. The high expression of Ki67 antigen, considered a poor prognostic, highlighted the highly proliferative nature of ATL cells [71].

Twenty-three patients in our series (table III) had a different phenotype. As described by others [72-75], these patients had a survival lower than that of typical cases (figure 5). Otherwise, double-negative cases, double-positive cases, cases with CD3-, and one with single intra-cytoplasmic expression of CD3 reflected the different stages of thymic ontogeny and may thus be interpreted as thymic progenitors (postulated normal counterpart). In this context, the weakness of expression of the CD3/TCR complex, always observed in ATL cells of our series and others [76,77], suggests that the target cell infected with HTLV-1 acquires an intermediate level of differentiation (CD7+ CD2 + CD3 / TCR low, level DP) between cortical immature thymocytes (CD7+ CD2 + CD3/TCR-) and mature thymocytes (CD2 + CD3/TCR +) [78]. The low expression of CD3/TCR could be attributable to the HTLV- 1/TAX protein which can *in vitro* reduce the transcription of pre-TCR alpha gene in human immature thymocytes [79,80]. Otherwise, CD7 is functionally associated with the CD3/TCR complex and acquired very early during thymic ontogeny [77,78]. So, the loss of CD7 could be a secondary step towards the completed ATL cell, but not compulsory, as suggested by our patients with CD7 + phenotype.

## Conclusion

We studied the epidemiologic, clinical, and biological characteristics of patients with ATL disease during 30 years of follow-up in Martinique. Our cohort is one of the largest non-Japanese cohorts of ATL. Our cases are similar to those described notably in Jamaica and Japan. The median survival time of our series confirmed the seriousness of this hematologic malignancy. The high frequency of skin lesions, especially in indolent forms, and their favorable prognostic meaning suggested that their early treatment would delay the expansion of ATL cells to other tissues. Our study confirmed the high frequency and the diversity of chromosomal abnormalities, not related to clinical aspects. The low expression of the CD3/TCR complex and the atypical phenotypes drive our focus to the origin of the HTLV-1 cell, pre-ATL.

## Data Availability

I declare the disponibility of all data referred to in the manuscript

## Acknowledgements

We thank Line Timard, Patricia Ulric-Gervaise and Nathalie Auguiac Volny-Anne for excellent work in flux cytometry.

## Bibliography

1- Uchiyama T, Yodoi J, Sagawa K, Takatsuki K, Uchino H. Adult T-cell leukemia: clinical and hematologic features of 16 cases. Blood 1977;50:481–92.

2- Poiesz BJ, Ruscetti FW, Gazdar AF, Burn PA, Minna JD, Gallo RC. Detection and isolation of type C retrovirus particles from fresh and cultured lymphocytes of a patient with cutaneous T-cell lymphoma. Proc Natl AcadSci USA 1980;77(12):7415–19.

3- Poiesz BJ, Ruscetti FW, Reitz MS, Kalyanaraman VS, Gallo RC. Isolation of a new type C retrovirus (HTLV) in primary uncultured cells of a patient with Sezary T-cell leukemia. Nature 1981;294:268–71.

4- Gessain A, Barin F, Vernant JC, Gout O, Maurs L, Calender A et al. Antibodies to human T-lymphotropic virus type-1 in patients with tropical spastic paraparesis. Lancet 1985;2(8452):407–10.

5- Rodgers-Johnson P, Gajdusek DC, Morgan OS, Zaninovic V, Sarin PS, Graham DS. HTLV-1 and HTLV-III antibodies and tropical spastic paraparesis. Lancet 1985 Nov 30;2(8466):1247–8.

6- Osame M, Usuku K, Izumo S, Ijichi N, Amitani H, Igata A et al. HTLV-1 associated myelopathy, a new clinical entity. Lancet 1986;1(8488):1031–2.

7- LaGrenade L, Hanchard B, Fletcher V, Cranston B, Blattner W. Infective dermatitis of Jamaican children: a marker for HTLV-I infection. Lancet 1990;336(8727):1345–7.

8- Bittencourt AL, Primo J, Oliveira MF. Manifestations of the human T-cell lymphotropic virus type 1 infection in childhood and adolescence. J Pediatr (Rio J) 2006;82(6):411–20.

9- Shimoyama M. Diagnostic criteria and classification of clinical subtypes of adult T-cell leukemia-lymphoma. A report from the Lymphoma Study Group (1984-87). Br J Haematol 1991;79(3):428–37.

10- Kamada N, Sakurai M, Miyamoto K, Sanada I, Sadamori N, Fukuhara S et al. Chromosome abnormalities in adult T-cell leukemia/lymphoma: a karyotype review committee report. Cancer Res 1992;52(6):1481–93.

11- Yamaguchi K, Matutes E, Catowsky D, Galton DAG, Nakada K, Takatsuki K. Strongyloïdes stercoralis as candidate co-factor for HTLV-1-induced leukaemogenesis. Lancet 1987 July 11;2(8550):94–5.

12- Plumelle Y, Gonin C, Edouard A, Buchet B, Thomas L, Brebion A et al. Effect of Strongyloïdes stercoralis infection and eosinophilia on age at onset and prognosis of adult T-cell leukemia. Am J Clin Pathol 1997;107:81–7.

13- Gabet AS, Mortreux F, Talarmin A, Plumelle Y, Leclercq I, Leroy A et al. High circulating proviral load with oligoclonal expansion of HTLV-1 bearing T cells in HTLV-1 carriers with strongyloidiasis. Oncogene 2000;19:4954–60.

14- Gillet NA, Cook LB, Laydon KDJ, Hlela C, Verdonck K, Alvarez C et al. Strongyloidiasis and infective dermatitis alter Human T Lymphotropic Virus-1 clonality in vivo. M. PLOS pathogens 2013;9(4):e1003263.

15- Schaffar-Deshayes L, Chavance M, Monplaisir N, Courouce AM, Gessain A, Blesonski S et al Antibodies to HTLV-1 p24 in sera of blood donors elderly people and patients with hemopoietic diseases in France and in French West indies. Int J Cancer 1984;34:667–70.

16- Monplaisir N, Valette Y, Desaphy Y, Neisson-Vernant C. Blood transfusion and HTLV-1 infection in Martinique. Editors, C Roma, JC Vernant, M Osame. In Neurology and Neurobiology: HTLV-1 and the nervous system. New York: Alan R. Liss;1989. p933.

17- Gessain A, Plumelle Y, Sanhadji K, Barin F, Gazzolo L, Constant-Desportes M et al. [Adult T-cell leukemia/lymphoma associated with HTLV-1 virus in Martinique: about of two cases]. Nouv Rev Fr Hematol 1986;28(2):107–13.

18- Gessain A, Cassar O. Epidemiological aspects and world distribution of HTLV-1 infection. Frontiers in Microbiology 2012;3(388):1–23.

19- Watanabe T. Current status of HTLV-1 infection. Int J Hematol 2011;94:430–4.

20- Plumelle Y, Pascaline N, Nguyen D, Panelatti G, Jouannelle A, Jouault H et al. Adult T-cell leukemia-lymphoma: a clinico-pathologic study of twenty-six patients from Martinique. Hematol Pathol 1993;7(4):251–62.

21- Satake M, Yamada Y, Atogami S, Yamaguchi K. The incidence of adult T-cell leukemia/lymphoma among human T-lymphotopic virus type 1 carriers in Japan. Leuk Lymphoma 2015;56(6):1806–12,doi:10.3109/10428194.2014.964700.

22- Plumelle Y, Michel S, Banydeen R, Delaunay C, Panelatti G. Characteristics of adult T-cell leukemia / lymphoma patients with long survival: Prognostic significance of skin lesions and possible beneficial role of valproic acid. Leukemia Res and Treatment 2015;http://dx.doi.org/10.1155/2015/476805.

23- Besson C, Gonin C, Brebion A, Delaunay C, Panelatti G, Plumelle Y. Incidence of hematological malignancies in Martinique, French West Indies, overrepresentation of multiple myeloma and adult T cell leukemia/lymphoma. Leukemia 2001;15:828–31.

24- Panelatti G, Plumelle Y, Arfi S, Pascaline N, Caplanne D, Jean-Baptiste G. [Sarcoidosis and leukemia/T-cell lymphoma associated with HTLV-1 virus infection in adult (a propos of a case]. Rev Med Intern 1992;299–301.

25- Merle H, Donnio A, Gonin C, Albert JC, Panelatti G, Plumelle Y. Retinal vasculitis caused by adult T-cell leukemia/lymphoma. Jpn J Ophtalmol 2005;49:41–5.

26- Tajima K. The 4th nation-wide study of adult T-cell leukemia/lymphoma (ATL) in Japan: estimates of risk of ATL and its geographical and clinical features. The T- and B-cell malignancy study group and co-authors. Int J Cancer 1990;45:237–43.

27- Gibbs WN, Lofters WS, Campbell M, Hanchard B, LaGrenade L, Cranston B et al. Non-Hodgkin lymphoma in Jamaica and its relation to adult T-cell leukemia-lymphoma. Annals Int Medicine 1987;106:361–8.

28- Bittencourt AL, Barbosa HS, Requiao C, da Sylva AC, Vandamme AM, Van Weyenberg J et al. An exceptionnal pediatric case of ATL, with a mixed CD4+ and CD8+ phenotype and a particulary indolent course. J Clin Oncol 2007;25:2480–2.

29- Senba M, Kawai K, Mori N. Pathogenesis of Metastatic Calcification and Acute Pancreatitis in Adult T-Cell Leukemia under Hypercalcemic State. Leuk Res Treatment 2012;2012:128617. doi:10.1155/2012/128617. Epub 2011 Dec 1.

30- Setoyama M, Katahira Y, Kanzaki T. Clinicopathologic analysis of 124 cases of Adult T-cell leukemia/lymphoma with cutaneous manifestations: the smouldering type with skin manifestations has a poorer prognosis than previously thought. J Dermatol 1999;26(12):785–90.

31- Yamaguchi T, Ohshima K, Karube K, Tutiya T, Kawano R, Suefuji H et al. Clinicopathological features of cutaneous lesions of adult T-cell leukemia/lymphoma. British J Dermatol 2005;152:76–81.

32- Adult T-cell leukemia/lymphoma in Bahia, Brazil: Analysis of prognostic factors in a group of 70 patients. Bittencourt AL, Das Gracas Vieira M, Brites CR, Farré L, Barbosa HS. Am J Clin Pathol 2007;128:875–82.

33- Dosaka N, Tanaka T, Miyashi Y, Imamura S, Kakizuca A. Examination of HTLV-I integration in the skin lesions of adult T-cell leukemia (ATL): independence of cutaneous-type ATL confirmed by Southern blot analysis. J Invest Dermatol 1991;96 :196–200.

34- Amano M, Kurokawa M, Ogata K, Itoh H, Kataoka H, Setoyama M. New entity, definition and diagnostic criteria of cutaneous adult T-cell leukemia/lymphoma: human T-virus type 1 proviral DNA load can distinguish between cutaneous and smoldering types. J Dermatol 2008;35(5):270–5.

35- Tsukasaki K, Tsushima H, Yamamura M, Hata T, Maeda T, Atogami S et al. Integration patterns of HTLV-1 provirus in relation to the clinical course of ATL: frequent clonal change at crisis from indolent disease. Blood 1997 Feb 1;89(3):948–56.

36- Bangham CRM, Cook Lucy B, Melaned A. HTLV-1 clonality in adult T-cell leukemia and non-malignant HTLV-1 infection. Semin Cancer Biol 2014;26(100):89–98.

37- Hanchard B, La Grenade L, Carberry C, Fletcher V, Williams E, Cranston B et al. Childhood infective dermatitis evolving into adult T-cell leukemia after 17 years. Lancet 1991;338 :1593–4.

38- Farre L, Paim de Oliviera MF, Primo J, Vandamme AM, Van Weyenberg J, Bittencourt AL Early sequential development of infective dermatitis, Human T Cell Lymphotropic virus type 1-associated myelopathy, and adult T-cell leukemia/lymphoma. Clin Infect Disease 2008;46:440–2.

39- Kohno T, Uchida H, Inomata H, Fukushima S, Takeshita M, Kikuchi M. Ocular manifestations of adult T-cell leukemia/lymphoma. Ophthalmology. 1993;100:1794–9.

40- Shibata K, Shimamoto Y, Nishimura T, Okinami S, Yamada H, Miyahara M. Ocular manifestations in adult T-cell leukemia/lymphoma. Ann Hematol 1997;74:163–8.

41- Merle H, Hage R, Meniane JC, Deligny C, Plumelle Y, Donnio A, Jean-Charles A. Retinal manifestations in adult T-cell leukemia/lymphoma related to infection by the human T-cell lymphotropic virus type 1. Retina 2016;36(7):364–71. doi.10.1097.

42- Kikuchi K. Development of thymic tumor in HTLV-1 transgenic rats under control of a lymphoid tissue specific promoter. Hokkaido IgakuZasshi 1997;72(5):545–64.

43- Kikuchi K, Ikeda H, Tsuchikawa T, Tsuji T, Tanaka S, Fugo K et al. A novel animal model of thymic tumor: development of epithelial thymoma in transgenic rats carrying human T lymphocyte virus type I pX gene. Int J Exp Pathol 2002;83:247–55.

44- Yoshioka R, Yamaguchi K, Yoshinaga T, Takatsuki K. Pulmonary complications in patients with adult T-cell leukemia. Cancer 1985;55:2491–4.

45- Bellon M, Lepelletier Y, Hermine O, Nicot C. Deregulation of microRNA involved in hematopoiesis and the immune response in HTLV-1 adult T-cell leukemia. Blood 2009;113(20):4914–7.

46- Nagafuji K, Harada M, Teshima T, Eto T, Takamatsu Y, Okamura T et al. Hematopoietic progenitor cells from patients with adult T-cell leukemia-lymphoma are not infected with human T-cell leukemia virus type 1. Blood 1993;82(9):2823–8.

47- Nimer SD, Gasson JC, Hu K, Smalberg I, Williams JL, Chen IS et al. Activation of the GM-CSF promoter by HTLV-I and HTLV-II tax proteins. Oncogene 1989;4(6):671–6.

48- Uchijima M, Sato H, Fujii M, Seiki M Tax proteins of Human-cell leukemia virus type 1 and 2 induce expression of the gene encoding erythroid-potentiating activity (Tissue Inhibitor of Metalloproteinases-1, TIMP-1). J Biol Chem 1994;269(21):14946–50.

49- Fazi F, Rosa A, Fatica A, Gelmetti V,De Marchis ML, Nervi C et al. A minicircuitry comprised of microRNA-223 and transcription factors NFI-A and C/EBP alpha regulates human granulopoiesis. Cell 2005;123:819–31.

50- Ohshima K, Kikuchi M, Yoshida T, Masuda Y, Kimura N. Lymph nodes in incipient adult T-cell leukemia lymphoma with Hodgkin’s disease-like histologic features. Cancer 1991;67:1622–8.

51- Santos JB1, Farré L, Batista Eda S, Santos HH, Vieira MD, Bittencourt AL The importance of flower cells for the early diagnosis of acute adult T-cell leukemia/lymphoma with skin involvement. Acta Oncol 2010;49(2):265–7. doi: 10.3109/02841860903428192.

52- Bazarbachi A, Plumelle Y, Carlos-Ramos J, Tortevoye P, Otrock Z, Taylor G et al. Meta-analysis on the use of zidovudine and interferon-alpha in adult T-cell leukemia/lymphoma showing improved survival in the leukemic subtypes. J Clin Oncol 2010;28(27):4177–83.

53- Hodson A, Crichton S, Montoto S, Mir N, Matutes E, Cwynarski K et al. Use of zidovudine and interferon alfa with chemotherapy improves survival in both acute and lymphoma subtypes of adult T-cell leukaemia/lymphoma. J Clin Oncol 2011;29(35):4696–701.

54- Takasaki Y, Iwanaga M, Imaizumi Y, Tawara M, Joh T, Kohno T et al. Long-term study of indolent adult T-cell leukaemia-lymphoma. Blood 2010;115(22):4337–43.

55- Cavrois M, Wain-Hobson S, Gessain A, Plumelle Y, Wattel E. Adult T-cell leukemia/lymphoma on a background of clonally expanding human T-cell leukemia virus type-1 positive. Blood 1996;88(12):4646–50.

56- Imaizumi Y, Iwanaga M, Tsukasaki K, Hata T, Tomonaga M, Ikeda S Natural course of HTLV-1 carriers with monoclonal proliferation of T lymphocytes (“pre ATL”) in a 20-year follow-up study. Blood 2005;105(2):903–4.

57- Tsukasaki K, Ikeda S, Murata K, Maeda T, Atogami S, Sohda H et al. Characteristics of chemotherapy-induced clinical remission in long survivors with aggressive adult T-cell leukemia/lymphoma. Leuk Res 1993;17(2):157–66.

58- Tsukasaki K, Hermine O, Bazarbachi A, Ratner L, Ramos JC, Harrington W JR et al. Definition, prognostic factors, treatment and response criteria of adult T-cell leukemia-lymphoma. A proposal from an international consensus meeting. J Clin Oncol 2009;27(3):453–9. Doi.101200/JC02008.18.2428.

59- Oshiro A, Tagawa H, Ohshima K, Karube B, Uike M et al. Identification of subtype-specific genomic alterations in aggressive adult T-cell leukemia/lymphoma. Blood 2006;107(11);4500–7.

60- Itoyama T, Chaganti RSK, Yamada Y, Tsukasaki K, Atogami S, Nakamura H et al. Cytogenetic analysis and clinical significance in adult T-cell leukemia/lymphoma: a study of 50 cases from the human T-cell leukemia virus type-1 endemic area, Nagasaki. Blood 2001;97(11):3612–20.

61- Miyamoto K, Sato J, Kitajima K, Togawa A, Suemaru S, Sanada H et al. Adult T-cell leukemia. Chromosome analysis of 15 cases. Cancer 1983;52(3):471–8.

62- Sadamori N, Nishino K, Kusano M, Tomonaga Y, Tagawa M, Yao E et al. Significance of chromosome 14 anomaly at band 14q11 in Japanese patients with adult T-cell leukemia. Cancer 1986;58(10);2244–50.

63- Fujita K, Yamasaki Y, Sawada H, Izumi Y, Fukuhara S, Uchino H. Cytogenetic studies on the adult T-cell leukemia in Japan. Leuk Res 1989;13(7):535–43.

64- Vermeer MH, van Doorn R, Dijkman R, Mao X, Whittaker S, van Voorst Vader PC et al. Novel and highly recurrent chromosomal alterations in Sézary syndrome. Cancer Res 2008;68(8):2689–98. doi: 10.1158/0008-5472.CAN-07-6398.

65- Mao X, Chaplin T, Young BD. Integrated genomic analysis of Sézary syndrome. Genet Res Int 2011;2011:980150. doi: 10.4061/2011/980150. Epub 2011 Nov 24.

66- Espinet B, Salgado R. Mycosis fungoïdes and Sézary syndrome. Methods Mol Biol 2013;973:175-88. doi:10.1007/978-1-62703-281-011.

67- Fukuda RI, Hayashi A, Utsunomiya A, Nukada Y, Fukui R, Itoh K et al. Alteration of phosphatidylinositol 3-kinase cascade in the multilobulated nuclear formation of adult T-cell leukemia/lymphoma (ATLL). Proc Natl Acad Sci USA 2005;102:15213–8.

68- Nakahata S, Ichikawa T, Maneesaay P, Saito Y, Nagai K, Tamura T et al. Loss of NDRG2 expression activates PI3K-AKT signalling via PTEN phosphorylation in ATLL and others cancers. Nat commun 2014 Feb 26;5:3393.doi:10.1038/ncomms4393.

69- Shirono K, Hattori T, Hata H, Nishimura H, Takatsuki K. Profiles of expression of activated cell antigens on peripheral blood and lymph node cells from different clinical stages of adult T-cell leukemia. Blood 1989 May 1;73(6):1664–71.

70- Berkawitz JL, Janik JE, Stewart DM, Jaffe ES, Stetler-Stevenson M, Shih JH et al. Safety, efficacy and pharmakocinetics/pharmacodynamics of daclizumab (anti CD25) in patients with adult T-cell leukemia/lymphoma. Clin Immunol 2014;155(2):176–87. doi:10.1016/J.Clinm.2014.09.012.

71- Yamada Y, Murata K, Kamihira S, Atogami S, Tsukasaki K, Sohda H et al. Pronostic significance of the proportion of Ki-67-positive cells in adult T-cell leukemia. Cancer 1991;67:2605–9.

72- Yamada Y, Kamihira S, Amagasaki T, Kinoshita K, Kusano M, Chiyoda S et al. Adult T cell leukemia with atypical surface phenotypes: clinical correlation. J Clin Oncol 1985;3:782–8.

73- Kamihira S, Sohda H, Atogami S, Toriya K, Yamada Y, Tsukasaki K et al. Phenotype diversity and prognosis of adult T-cell leukemia. Leukemia Res 1992;16(5):435–41.

74- Kamihira S, Sohda H, Atogami S, Fukushima T, Toriya K, Miyazaki Y et al. Unusual morphological features of adult T-cell leukemia cells with aberrant immunophenotype. Leuk Lymphoma 1993;12:123–30.

75- Shimauchi T, Hirokawa Y, Tokura Y. Purpuric adult T-cell leukemia/lymphoma: expansion of unusual CD4/CD8 double-negative malignant T cells expressing CCR4 but bearing the cytotoxic molecule granzyme B. Br J Dermatol 2005;152:350–2.

76- Kobayashi S, Tian Y, Ohno N, Yuji K, Ishigaki T, Isobe M et al. -The CD3 versus CD7 plot in multicolor flow cytometry reflects progression of disease stage in patients infected with HTLV-1. PloS One 2013; 8(1):e53728.doi:10.1371/journal.pone.0053728.

77- Akl H, Badran B, Dobirta G, Manfouo-Foutsop G, Moshitta M, Merimi M et al. Progressive loss of CD3 expression after HTLV-1 infection results from chromatin remodelling affecting all the CD3 genes and persists despite early viral genes silencing. J Virol 2007;4:85.doi:10.1186/1743-422X-4-85.

78- Maguer-Satta V, Gazzolo L, Duc Dodon M. Human immature thymocytes as target cells of the leukemogenic activity of human T-cell leukemia virus type 1. Blood 1995;86(4):1444–52.

79- Wencker M, Sausse C, Derse D, Gazzolo L, Duc Dodon M. Human T-cell leukemia virus type 1 tax protein down-regulates pre-T-cell receptor alpha gene transcription in human immature thymocytes. J Virol 2007;81(1):301–8.

80- Villaudy J, Wencker M, Gadot N, Gillet NA, Scoazec JY, Gazzolo L et al. HTLV-1 propels thymic human T cell development in “human immune system” Rag2-/-gamma C-/-mice. PLOS pathogens 2011;7(9) e1002231.

